# Thematic analysis of a United Kingdom-wide survey to explore women’s perceptions and concerns about assisted reproductive technology

**DOI:** 10.1101/2025.07.16.25331627

**Authors:** Faiza Afzal, Ling Yin Fritz Wong, Mitana Purkayastha, Yan Lu, Philippa Rees, Melissa A. Richard, Carrie L. Williams, Philip J. Lupo, Barbara Luke, Alastair G. Sutcliffe

## Abstract

Over 300,000 children were born in the United Kingdom (UK) through assisted reproductive technology (ART). ART is associated with prematurity and congenital anomalies, while long-term health risks are currently considered low. Large-scale qualitative research exploring women’s perspectives on these issues remains limited. Therefore, this study investigates women’s concerns about health and educational outcomes following ART, and their views on information provision and national database studies.

Women who underwent ART, were considering ART or conceived naturally were invited to participate voluntarily. An anonymous online survey was distributed via social media channels of a UK-based fertility charity and accepted responses for 8 months. Free-text responses underwent thematic analysis, and descriptive statistics were produced for categorical responses.

Of the 562 participants, most were aged 25–40 (74.5%), White (93.2%) and held a degree or higher qualification (87.9%). The majority underwent ART through the private sector (42.3%) in the 2020s (69.1%) and had successful childbirth (52.4%). While up to 82.5% of respondents reported no health and educational concerns, others were concerned about reproductive, endocrine and hormone treatment-related cancer risks in themselves, in addition to potential reproductive, neurological and developmental issues in their offspring. Most participants (up to 91.9%) stated they received no information about ART-related health and educational outcomes in themselves and their child; those who did cited healthcare professionals or videos as sources. More than half (68.3%) supported using national databases to study children’s outcomes, and they expressed that such findings should be disseminated sensitively through healthcare professionals before treatment.

Although most women reported no concerns or information about long-term health and educational outcomes for themselves or their children, many expressed concerns. The lack of information and support for database studies can inform patient-centred communication that addresses women’s concerns. Future ART studies may consider incorporating patient and public involvement surveys to shape research priorities.

## Introduction

Over the last 40 years, assisted reproductive technology (ART), defined as ‘all treatments or procedures that include the in vitro handling of both human oocytes and sperm, or embryos, for the purpose of establishing a pregnancy’ (Zegers-Hochschild et al., 2009), has steadily increased, with approximately 12 million children born after ART worldwide (eClinicalMedicine, 2023). Over 300,000 children were born in the United Kingdom (UK) through ART, and individuals requiring ART to conceive are a growing group in the UK (Human Fertilisation and Embryology Authority, 2025). This being partially due to the prevailing social trend amongst couples towards delaying their first planned pregnancy, but could also reflect the impact of lifestyle factors such as obesity and underlying chronic health conditions (Glazer et al., 2017).

ART procedures include in vitro fertilisation (IVF), intra-cytoplasmic sperm injection (ICSI) and related micromanipulation techniques, but does not include intrauterine insemination. It has been shown that children born after ART have more hospital admissions than naturally conceived population controls (Sutcliffe et al., 2023). while several studies have focused on birth and childhood outcomes after undergoing ART (Helmerhorst et al., 2004), little has been done to specifically examine the effects of subfertility on these outcomes (Wisborg et al., 2010). Even less well-studied are the health outcomes of subfertile women who have received ART or the health outcomes of subfertile men. Subfertility, with or without ART, has previously been shown to be correlated with higher rates of: (a) prematurity, low birth weight (Wisborg et al., 2010) and congenital malformations in children, and hypertension in adolescence (Meister et al., 2018); (b) ovarian and breast cancer in women (Williams et al., 2018); and (c) testicular cancer (Ventimiglia et al., 2016) and potentially diabetes in men (Glazer et al., 2017) (although the evidence is limited).

The potential effects of ART on the short- and long-term health of these women and their children could be a source of concern for patients. In a survey conducted by our team in 2006, families with ART-conceived children reported potential general health risks to their children as their paramount concern (Fisher-Jeffes et al., 2006).

The aim of this thematic analysis was to explore the views of women who are considering fertility treatment, or those that have undergone fertility treatment. This was part of the preparation for a national data linkage study funded by Wellcome and the National Institutes of Health (NIH) called ‘United Kingdom Longitudinal Population Cohort Studies of Subfertile Individuals and Children Conceived after Fertility Treatments’ following the principle of nothing by us without us.

The objectives were to establish: health/educational outcome concerns, information provided by healthcare professionals (HCP) and perceptions of national database studies. The present study provides new insights on women’s concerns about their health outcomes and their ART-conceived child’s health and educational outcomes. It also offers insights on how information was presented to them at the fertility clinic, and their views on national database studies, including our own studies (https://liftresearchucl.com/, https://enchantresearchucl.com/).

## Methods

### Participants

Participants were identified by convenience (volunteer) sampling, where participants volunteered to take part in the survey. Women who had undergone or were considering fertility treatment were invited to participate in the survey.

### Survey design and distribution

The cross-sectional, English, online survey was designed and hosted on Qualtrics and distributed by the social media channels of Fertility Network UK, a charity which provides support for those with fertility issues across the UK. These included Instagram, Facebook and Twitter/X. No paid advertisements were used. The survey was anonymous and did not capture any personal identifiable data. The survey was active for 8 months.

The survey captured data across four domains (Table 1) to enable us to address our objectives. Domain 1 highlighted participant demographics (e.g., age, ethnicity, stage of treatment and type of clinic). Domain 2 explored participants’ concerns about their long-term health outcomes after fertility treatment and their ART-conceived child. Participants were also asked whether they were concerned about the educational outcomes of their ART-conceived child. For Domain 3, they were asked if any information about health outcomes for themselves or their children following fertility treatment was presented at the clinic. For Domain 4, participants expressed their views on national database studies and our group’s national data linkage study. All survey questions were optional.

**Table 1.** Survey questions and specific domains.

| Domain | Survey questions |
| --- | --- |
| 1. Experience with fertility treatment and demographics | Q1: Are you considering fertility treatment, have undergone fertility treatment or have conceived naturally?<br>Q2: Have you had children following assisted reproductive technology (ART)?<br>Q8: If you have undergone fertility treatment, how long ago was it?<br>Q9: Was it at a Private or NHS clinic?<br>Q15: What age group category applies to you?<br>Q16: What is your ethnic group?<br>Q17: What is your educational background? |
| 2. Concerns about health and educational outcomes | Q3: Do you have any particular concerns about the long-term health outcomes of your child/children? If yes, what are your concerns?<br>Q5: Do you have any particular concerns about the long-term educational outcomes of your child/children? If yes, what are your concerns?<br>Q10: Were potential short- and long-term health outcomes something you considered before/during/after fertility treatment?<br>Q11: Do you have any particular concerns regarding long-term health outcomes after fertility treatment? |
| 3. Concerns about information presented to women | Q4: How was the information on your ART child's health outcomes presented to you (if at all)?<br>Q6: How was the information on your child's educational outcomes presented to you (if at all)?<br>Q12: How was information on your long-term health outcomes (following ART) presented to you (if at all)? |
| 4. Perceptions of the methodology and dissemination of national database studies | Q7: What are your thoughts about analysing large anonymous data sets about your child's health and educational outcomes?<br>Q13: What are your thoughts about using large administrative patient data sets to retrieve information?<br>Q14: What are your views on how the findings of these studies could/should be presented to people undergoing fertility treatment? |

**Abbreviations:** ART, assisted reproductive technology; NHS, National Health Service;
Q, question number.

### Data analysis

Qualitative analysis of free-text responses (Q3–14) followed an inductive, post-positivist approach to Braun and Clarke’s (2006) thematic analysis. Each phase of this analysis was further guided by Nowell et al.’s (2017) approach to establish trustworthiness. LYFW manually coded all responses on NVivo 14 and connected the codes into broader themes and sub-themes. The resulting codebook was then independently reviewed by FA and discussed in regular meetings. This process was repeated five to seven times to iteratively refine the themes, with reflexive memos and audit trails documented throughout. Responses indicating ‘N/A’ were excluded from the analysis using pairwise deletion. The frequency of codes was counted to derive percentages for comparison across questions. Illustrative quotes were identified to support the data. Quantitative analysis of categorical responses (Q1, 2 and 15–17) was performed to produce descriptive statistics.

### Ethical approval and consent

Ethical approval was obtained from University College London’s Research Ethics Committee (28371/001). A disclaimer was added to the participant information sheet with contact details of Fertility Network UK. This was to ensure that participants were provided adequate support if they were concerned or affected by any of the questions in the survey. All participants provided informed consent to participate in the research.

## Results

Of the 1273 times the survey was accessed, 575 responses were submitted. Thirteen submissions provided empty answers to all questions and were excluded; 562 (44.1%) participants were included for analysis. As participation in each question was optional, the total number of responses varied across questions (mean = 454 responses). Key results from the four domains are presented below.

### Domain 1: Experience with fertility treatment and demographics (Q1, 2, 8, 9 and 15–17)

Most respondents were 25–40 years old (74.5%), White (93.2%) and held a degree or higher qualification (87.9%). They mostly underwent fertility treatment in the private sector in the 2020s and had successful childbirth (Table 2).

**Table 2.** Demographic and fertility treatment characteristics of respondents.

| **Characteristics** | **Responses (*n*)** | **%** |
| --- | --- | --- |
| **Age group (Q15)** |  |  |
| Under 25 | 1 | 0.2 |
| 25–40 | 407 | 74.5 |
| 40–60 | 138 | 25.3 |
| 60+ | 0 | 0.0 |
| **Total** | **546** | **100** |
| **Ethnic group (Q16)** |  |  |
| White | 507 | 93.2 |
| Asian/Asian British | 17 | 3.1 |
| Black/Black British/Caribbean/African | 9 | 1.7 |
| Mixed or multiple ethnic groups | 5 | 0.9 |
| Other ethnic group | 6 | 1.1 |
| **Total** | **544** | **100** |
| **Highest educational qualification (Q17)** |  |  |
| No formal qualifications | 3 | 0.6 |
| GCSE's or equivalent | 21 | 3.9 |
| A Levels or equivalent | 42 | 7.7 |
| Degree or equivalent | 274 | 50.5 |
| Higher degree or equivalent | 203 | 37.4 |
| **Total** | **543** | **100** |
| **Stage of fertility treatment (Q1)** |  |  |
| Undergone fertility treatment | 496 | 88.3 |
| Considering fertility treatment | 62 | 11.0 |
| Conceived naturally | 4 | 0.7 |
| **Total** | **562** | **100** |
| **Childbirth following ART (Q2)** |  |  |
| Yes | 294 | 52.4 |
| No | 267 | 47.6 |
| **Total** | **561** | **100** |
| **Year receiving fertility treatment (Q8)** |  |  |
| 1990–1999 | 3 | 0.5 |
| 2000–2009 | 13 | 2.3 |
| 2010–2019 | 150 | 26.6 |
| 2020–2024 | 389 | 69.1 |
| Not yet | 8 | 1.4 |
| **Total** | **563\*** | **100** |
| **Healthcare sector providing fertility treatment (Q9)** |  |  |
| Private | 213 | 42.3 |
| NHS | 179 | 35.5 |
| Both private and NHS | 112 | 22.2 |
| **Total** | **504** | **100** |

\* Some participants underwent more than one round of fertility treatment.
**Abbreviations:** ART, assisted reproductive technology; GCSE, General Certificate of Secondary Education; N/A, not applicable; NHS, National Health Service; Q, question number.

### Domain 2: Concerns about health and educational outcomes (Q3, 5, 10 and 11)

The themes in Domain 2 were categorised into three main sub-themes (Table 3). In relation to Q3, 5 and 11, over half of the respondents (66.9%, 82.5% and 51.8%, respectively) reported no concerns about long-term outcomes after fertility treatment: *‘No, at the moment I just want a child’* (P237); *‘Not really, it’s not something that has really been discussed with us’* (P339).

**Table 3.**
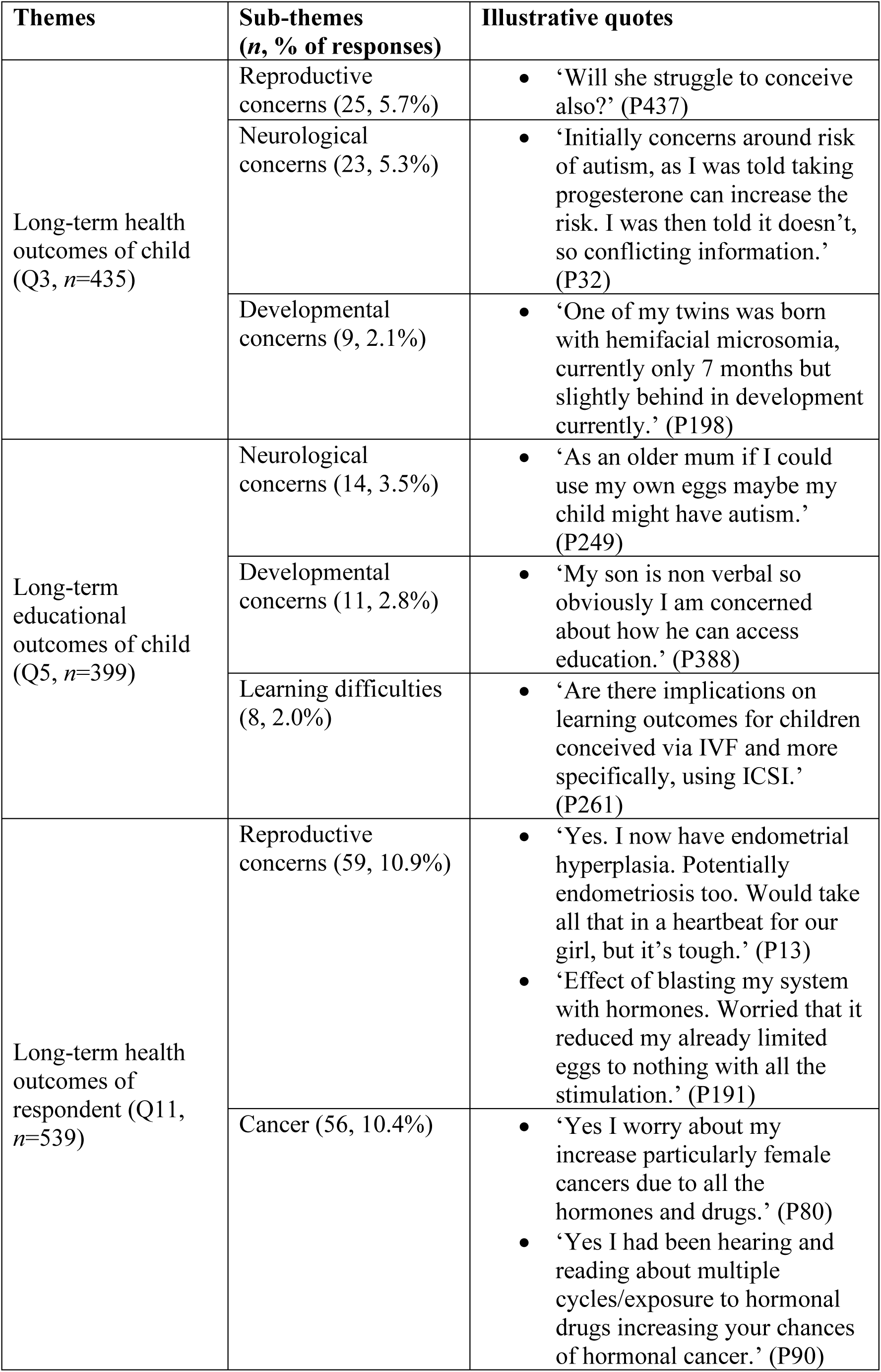
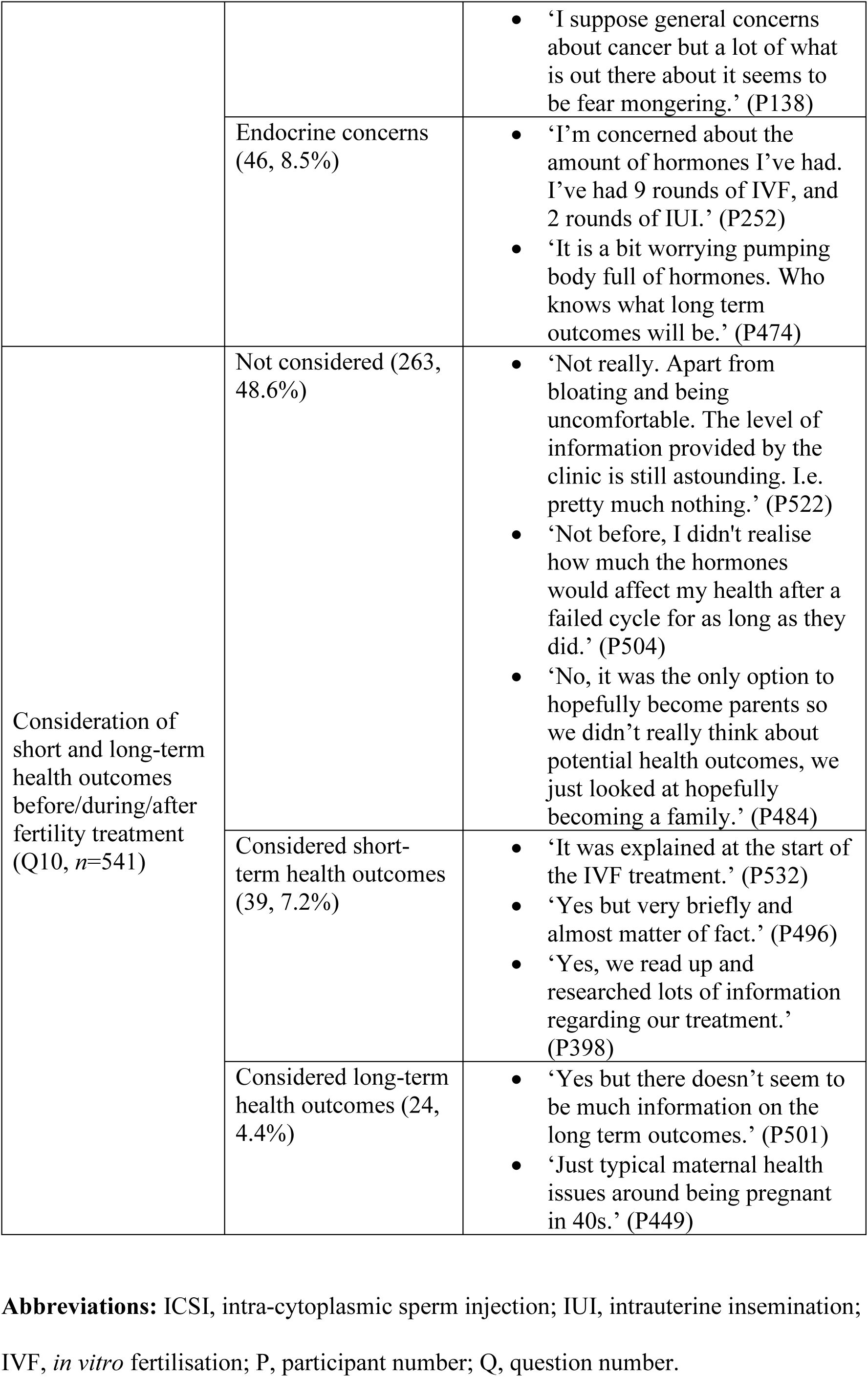
Concerns about health and educational outcomes.

Concerned individuals gave responses about the reproductive, neurological and developmental outcomes of their child born after fertility treatment: *‘They could possibly have the same fertility issues as us, their parents’* (P446). They also expressed specific concerns about educational outcomes of the child, including neurological concerns, developmental concerns and learning difficulties. Almost half (48.6%) said that they had not considered the long/short-term health outcomes of fertility treatment. More participants were concerned about health outcomes (33.1%–48.2%) than educational outcomes in their child/children (17.5%). The entire range of concerns expressed by participants in Domain 2 is shown in Figure 1. The three main sub-themes of Domain 1 were then further classified (Figure 2). The main reproductive concern mentioned was fertility issues (n=16, 3.6%) (Figure 2A). The main neurological concerns reported for long-term health and educational outcomes of the ART-conceived child were autism and attention deficit hyperactivity disorder (Figures 2A and 2B). Developmental concerns included developmental issues and developmental delays (Figure 2B). Of the participants that did not consider long/short-term health outcomes after fertility treatment, 14 indicated that the benefits outweighed the risks. Sixteen participants had health concerns after fertility treatment, eight participants during fertility treatment and six participants before fertility treatment (Figure 2C). The main reproductive concerns in women who were considering or undergoing ART were early menopause, fertility issues and endometriosis (Figure 2D). The main concerns about cancer were the association between hormones and cancer, cancer in general, breast cancer and ovarian cancer. Endocrine concerns included hormonal exposure and weight gain.

**Figure 1.**
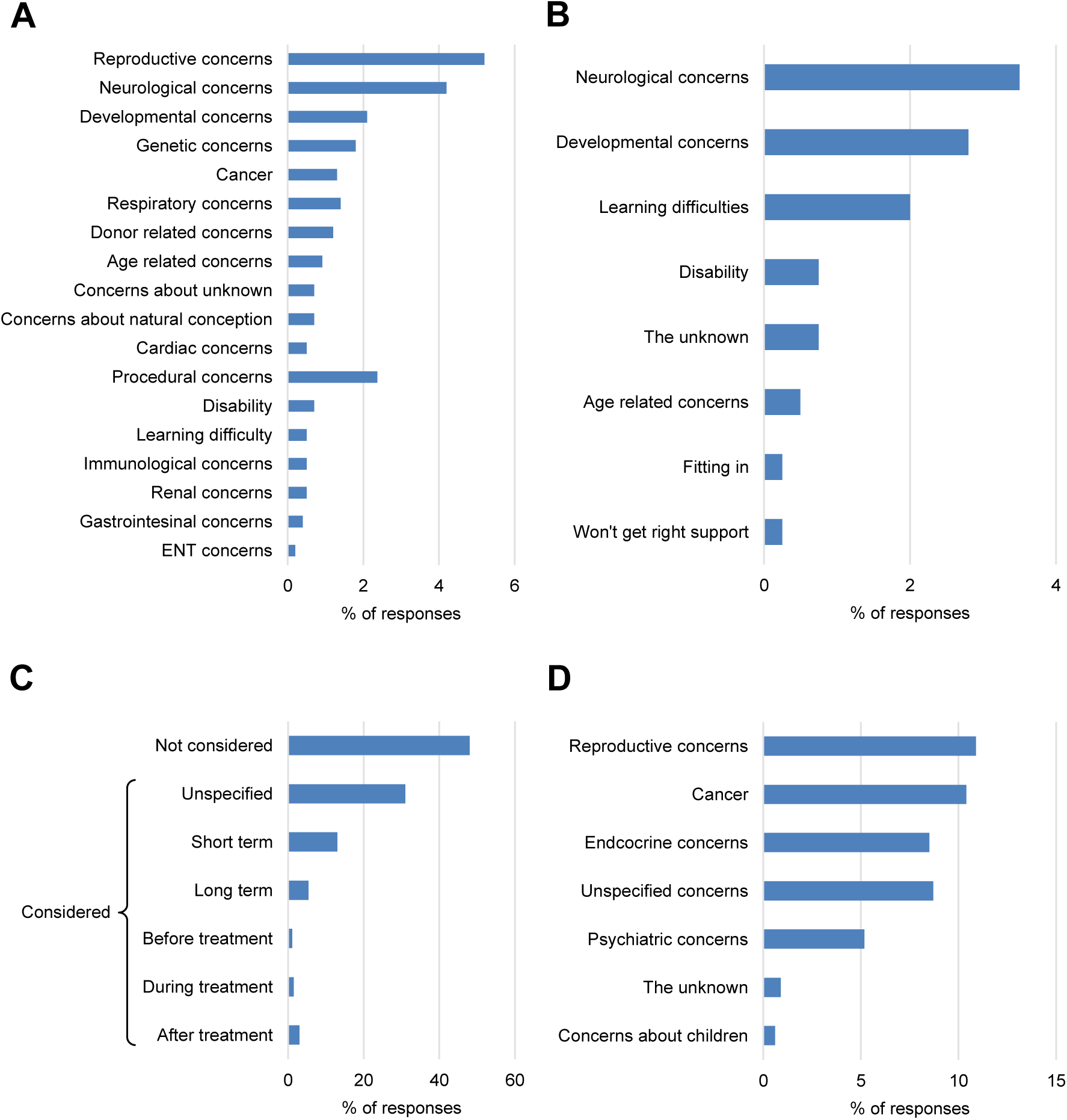
Concerns about (A) long-term health outcomes in child, (B) long-term educational outcomes in child, (C) consideration of long- and short-term health outcomes before, during and after fertility treatment and (D) long-term health outcomes in respondent. **Abbreviation:** ENT, ear, nose and throat.

**Figure 2.**
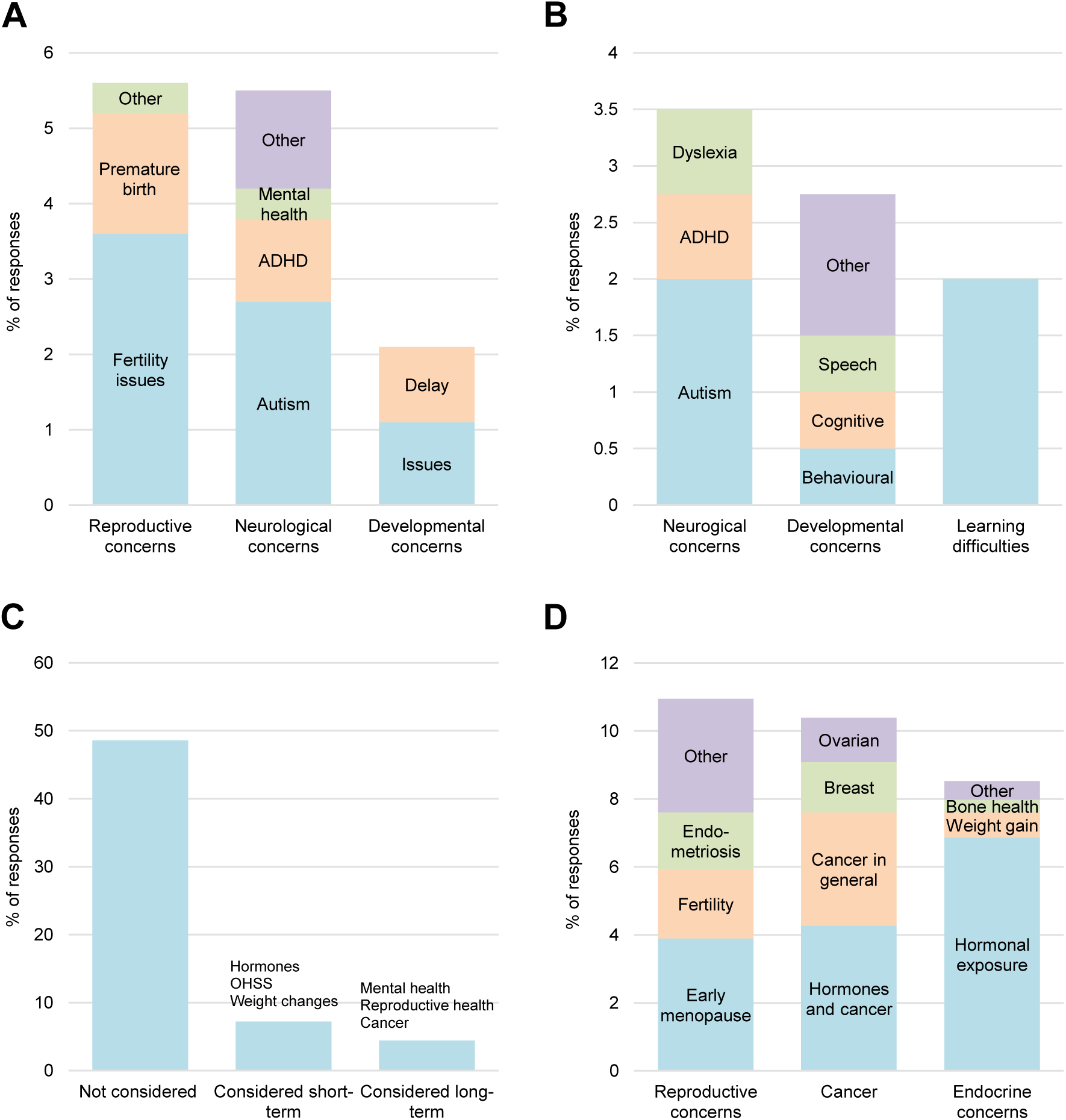
Top three concerns about (A) long-term health outcomes in child, (B) long-term educational outcomes in child, (C) consideration of long- and short-term health outcomes before, during and after fertility treatment and (D) long-term health outcomes in respondent. **Abbreviations:** ADHD, attention deficit hyperactivity disorder; OHSS, ovarian hyperstimulation syndrome.

### Domain 3: Concerns about information presented to women (Q4, 6 and 12)

Specific themes are shown in Table 4, and themes were classified into sub-themes (Figure 3). The majority of participants (75.2% in Q4 and 91.9% in Q6) said no information about their ART-conceived child’s health and educational outcomes were presented to them at the fertility clinic. Twenty-three participants (9.5%) reported that some information about their child’s health outcomes was provided by HCP and 3.3% said information was provided via videos (Figure 4A). Furthermore, 56.6% of participants said no information was provided to them about their health outcomes following fertility treatment (Figure 4A): *‘I have no idea what I should be worried about, my private fertility clinic never explained any risks or research findings with me’* (P412).

**Figure 3.**
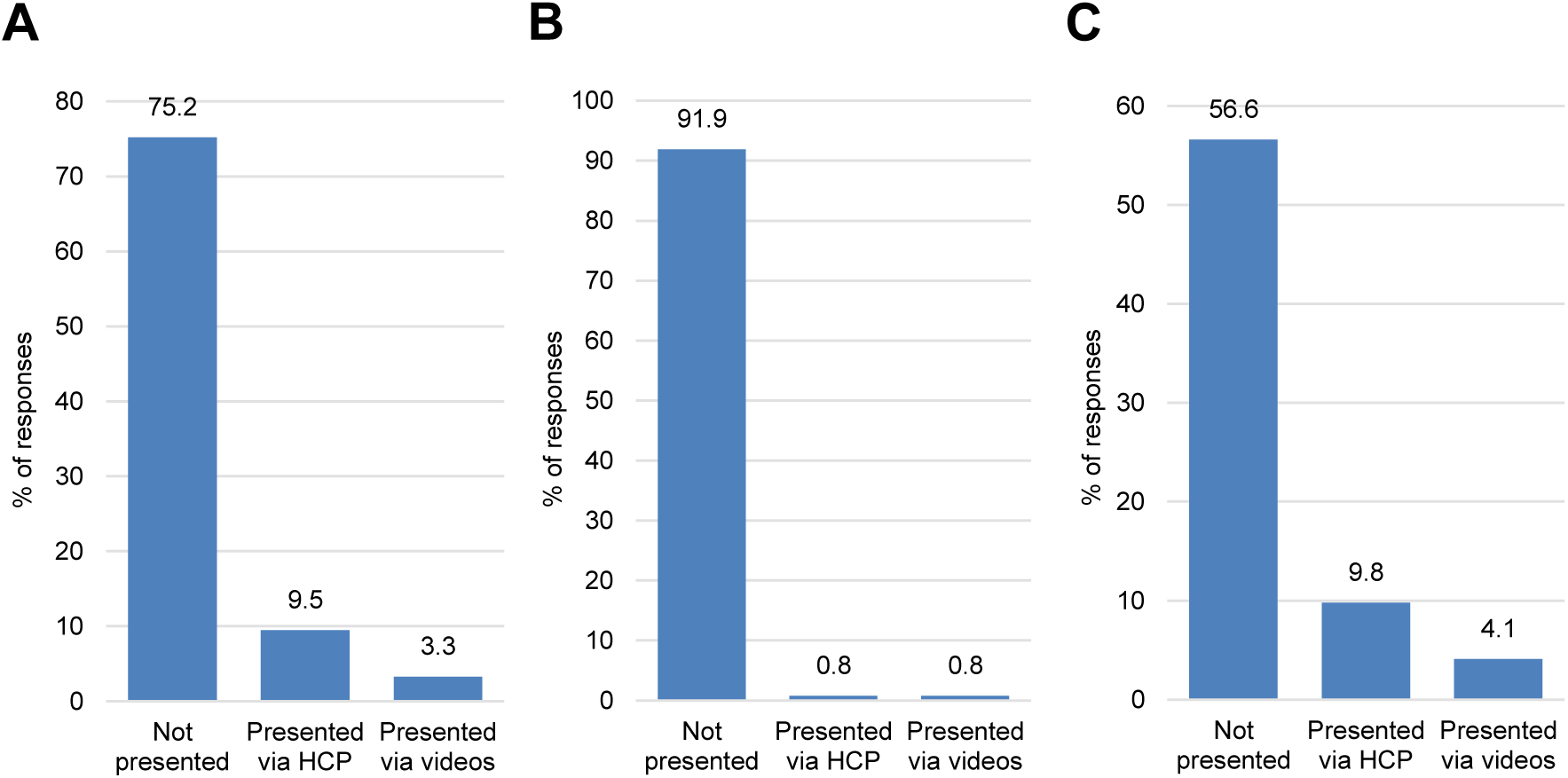
Presentation of information about (A) long-term health outcomes in ART-conceived child, (B) long-term educational outcomes in ART-conceived child and (C) long-term health outcomes of respondent. **Abbreviation:** HCP, healthcare professionals.

**Figure 4.**
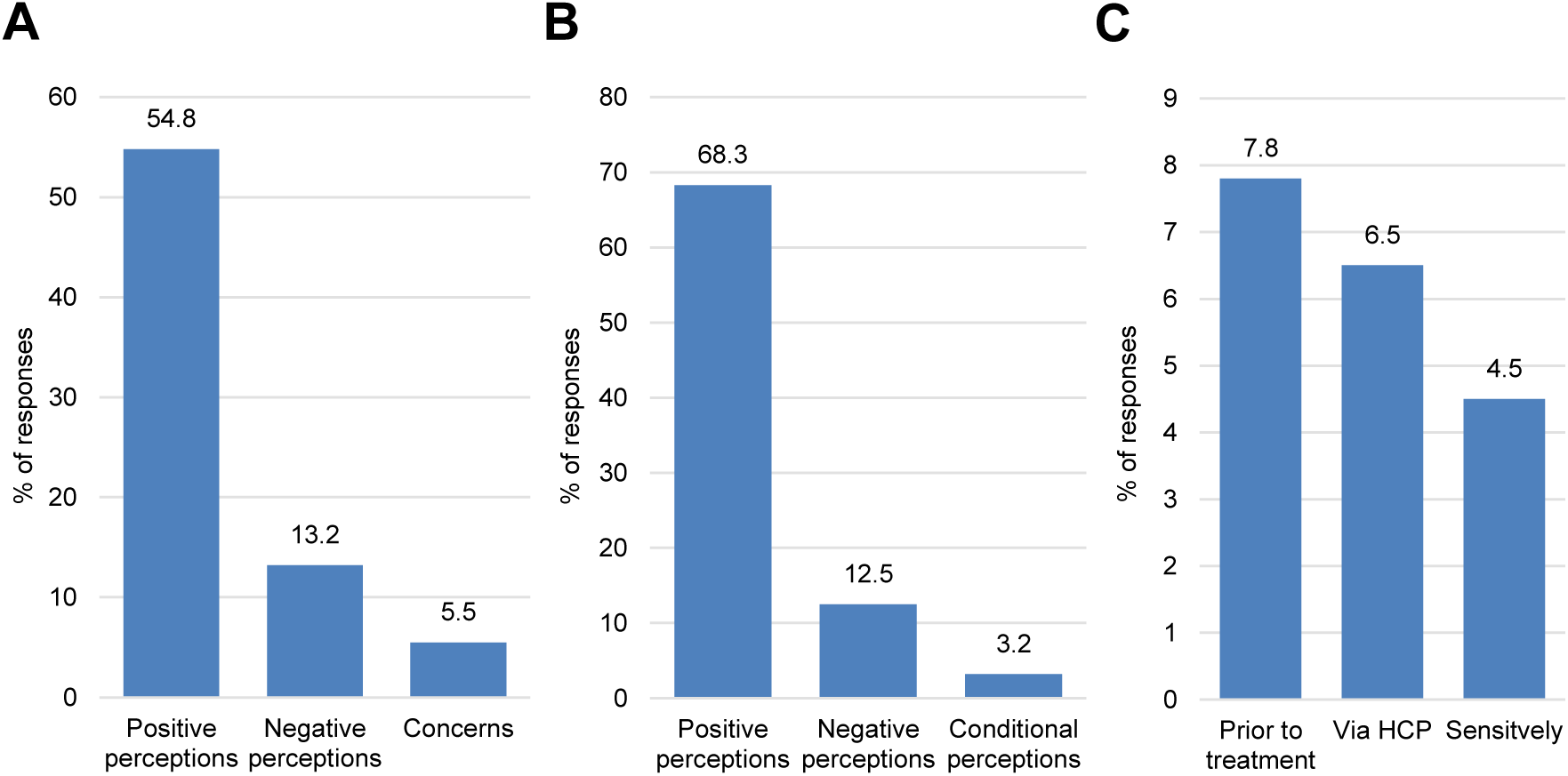
Perceptions of (A) study methodology: analysing data sets about health and educational outcomes, (B) study methodology: using large administrative patient data sets and (C) study dissemination. **Abbreviation:** HCP, healthcare professionals.

**Table 4.**
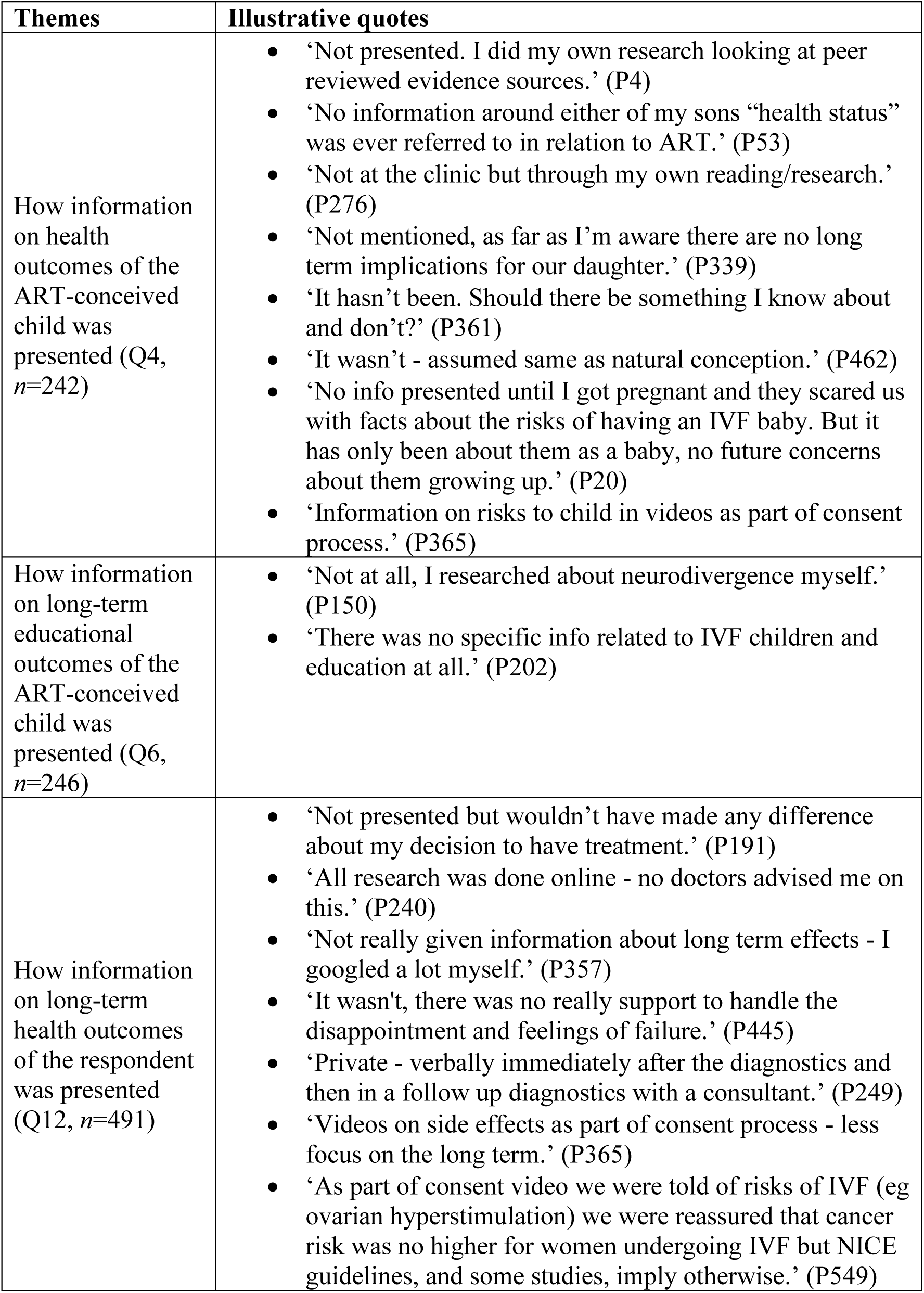
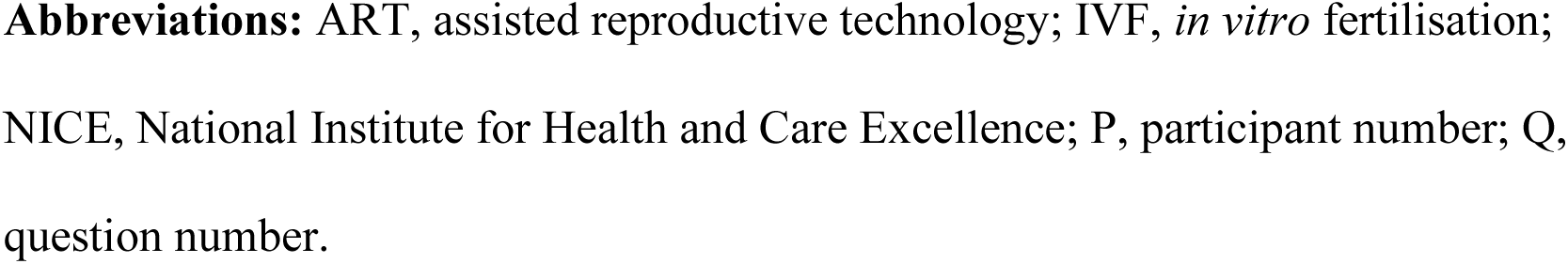
Concerns about how information on health and educational outcomes was presented to respondents.

| Themes | Illustrative quotes |
| --- | --- |
| How information on health outcomes of the ART-conceived child was presented (Q4, n=242) | • ‘Not presented. I did my own research looking at peer reviewed evidence sources.’ (P4)<br>• ‘No information around either of my sons “health status” was ever referred to in relation to ART.’ (P53)<br>• ‘Not at the clinic but through my own reading/research.’ (P276)<br>• ‘Not mentioned, as far as I’m aware there are no long term implications for our daughter.’ (P339)<br>• ‘It hasn’t been. Should there be something I know about and don’t?’ (P361)<br>• ‘It wasn’t - assumed same as natural conception.’ (P462)<br>• ‘No info presented until I got pregnant and they scared us with facts about the risks of having an IVF baby. But it has only been about them as a baby, no future concerns about them growing up.’ (P20)<br>• ‘Information on risks to child in videos as part of consent process.’ (P365) |
| How information on long-term educational outcomes of the ART-conceived child was presented (Q6, n=246) | • ‘Not at all, I researched about neurodivergence myself.’ (P150)<br>• ‘There was no specific info related to IVF children and education at all.’ (P202) |
| How information on long-term health outcomes of the respondent was presented (Q12, n=491) | • ‘Not presented but wouldn’t have made any difference about my decision to have treatment.’ (P191)<br>• ‘All research was done online - no doctors advised me on this.’ (P240)<br>• ‘Not really given information about long term effects - I googled a lot myself.’ (P357)<br>• ‘It wasn’t, there was no really support to handle the disappointment and feelings of failure.’ (P445)<br>• ‘Private - verbally immediately after the diagnostics and then in a follow up diagnostics with a consultant.’ (P249)<br>• ‘Videos on side effects as part of consent process - less focus on the long term.’ (P365)<br>• ‘As part of consent video we were told of risks of IVF (eg ovarian hyperstimulation) we were reassured that cancer risk was no higher for women undergoing IVF but NICE guidelines, and some studies, imply otherwise.’ (P549) |

**Abbreviations:** ART, assisted reproductive technology; IVF, *in vitro* fertilisation; NICE, National Institute for Health and Care Excellence; P, participant number; Q, question number.

### Domain 4: Perceptions of the methodology and dissemination of national database studies (Q7, 13 and 14)

Table 5 shows the specific themes and quotes from participants and these themes are further categorised into sub-themes (Figure 4). More than half (54.8%) had positive views about large national datasets to retrieve information about children’s health and educational outcomes: *‘I agree with the methodology. Analysing anonymous data is a good option to ensure privacy of patients’* (P1). Some had negative perceptions (13.2%) and 5.5% had concerns (Figure 4A). 68.3% of participants had positive perceptions about using large patient datasets to retrieve information: *‘Studies like these will help those considering fertility treatment in the future’* (P50). 12.5% had negative perceptions and 3.2% had conditional perceptions related to data confidentiality (Figure 4B). 7.8% said that study findings should be disseminated prior to treatment at the clinic. 6.5% said the results should be communicated by HCP across hospitals and clinics (Figure 4C): *‘People should be informed of potential risks when they have their first appointment with IVF clinic and further discussion should be offered’* (P86).

**Table 5.**
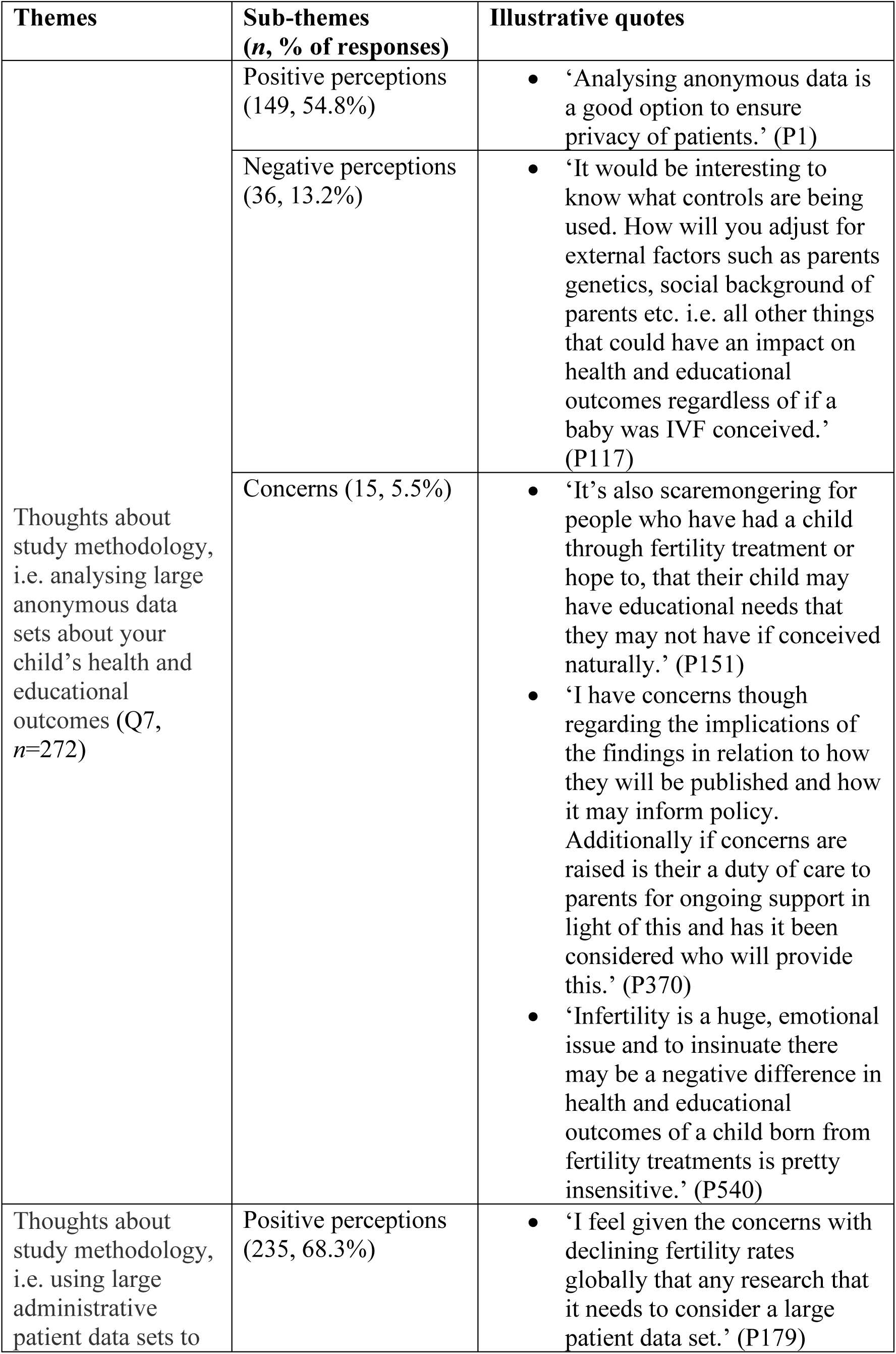
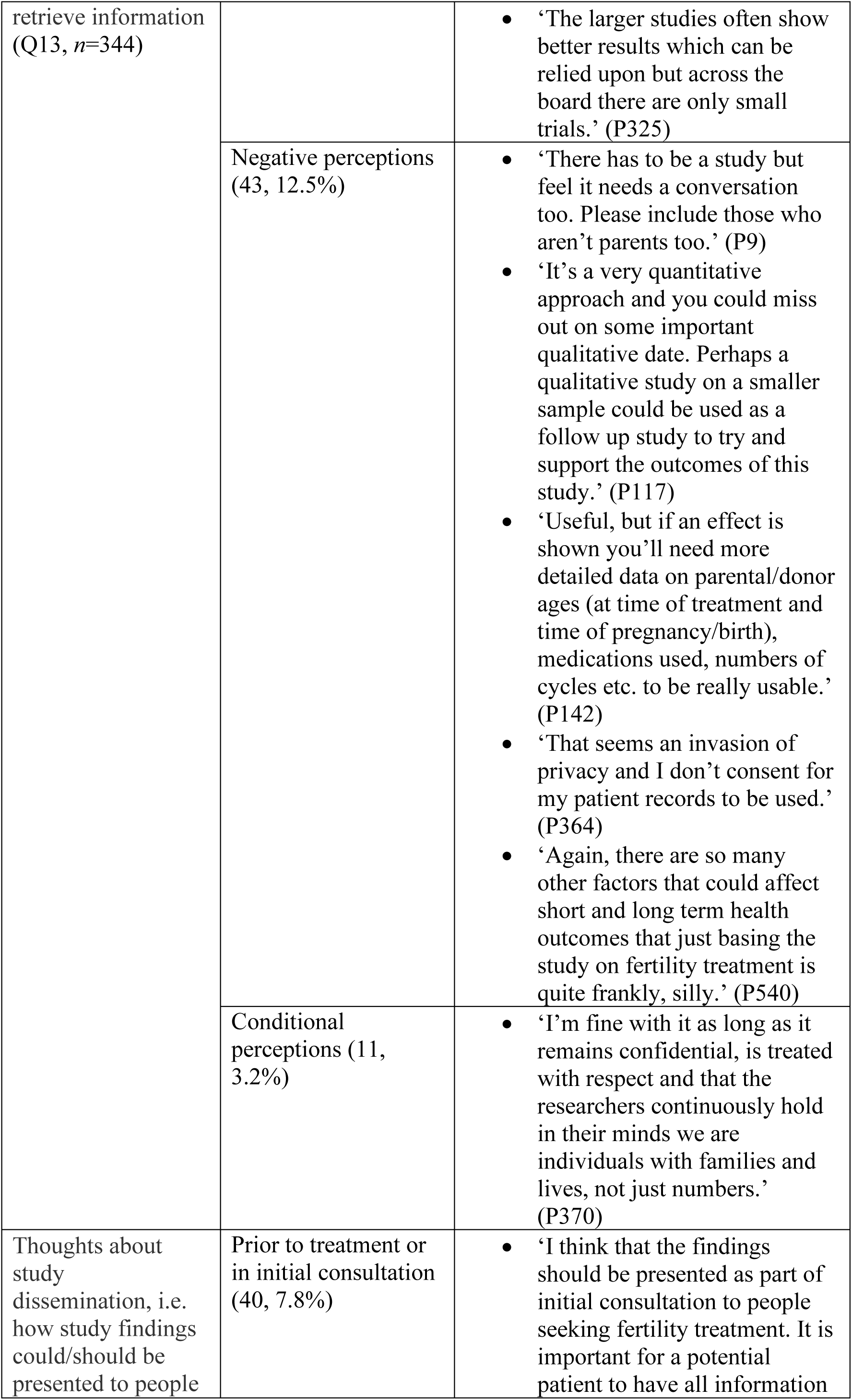
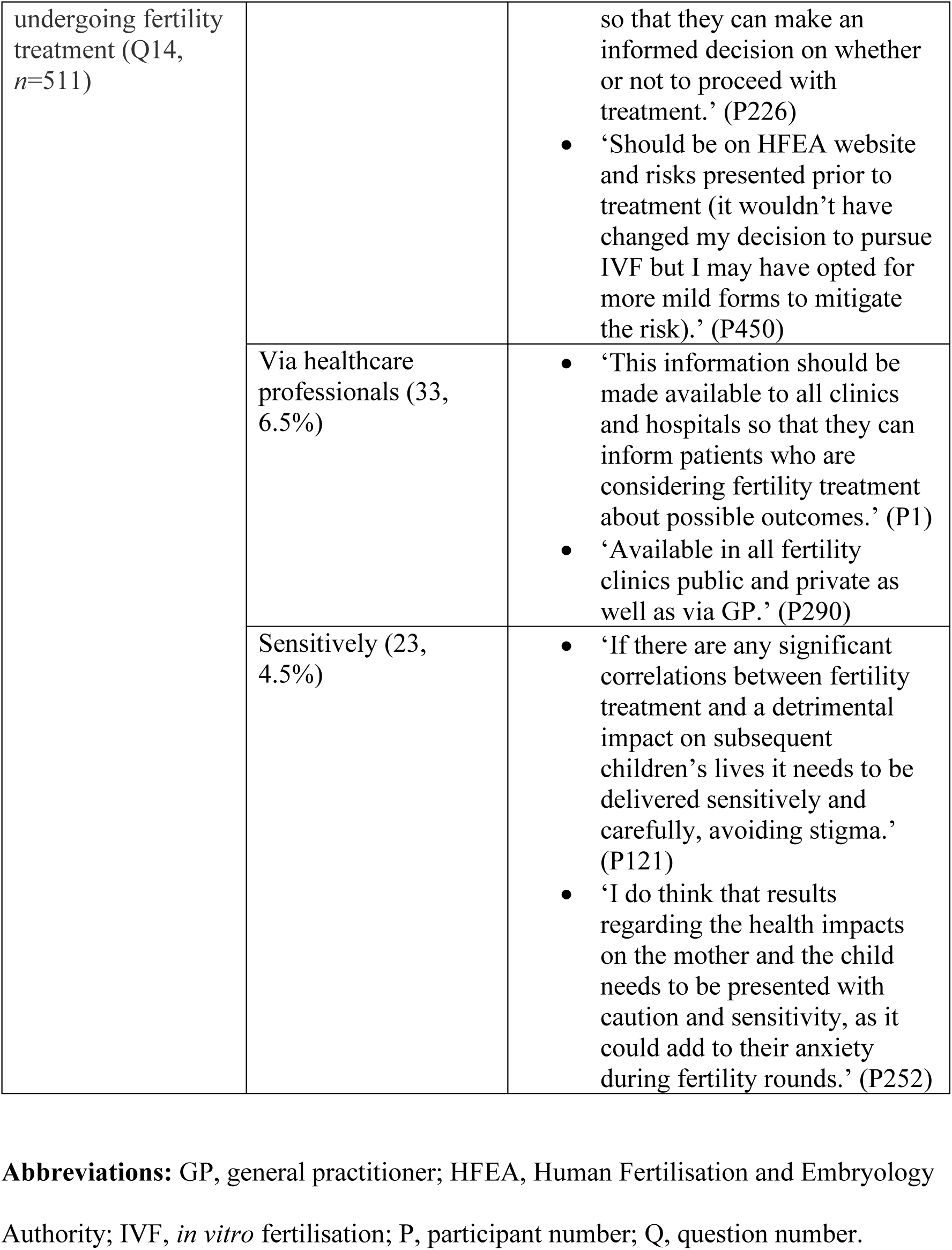
Perceptions of the methodology and dissemination of national database studies.

## Discussion

### Main findings

To our knowledge, this is the first large-scale qualitative survey to investigate the perceptions and concerns of women undergoing or considering ART. A key finding of this study is that most participants reported no concerns about health and educational outcomes for themselves and their ART-conceived child and interestingly, these women received no information on these issues from clinics. Receiving limited information from clinics could prevent women from recognising potential ART risks that may warrant concern.. Among those who expressed concerns, child health concerns focused on reproductive, neurological and developmental outcomes, while maternal concerns related to reproductive conditions, cancer risk and hormonal exposure. These findings highlight that, beyond achieving pregnancy, women undergoing or considering ART are attentive to the long-term implications of the treatment. This is often intertwined with the hope, desperation and unease they experienced during fertility treatment, as reflected in participants’ quotes. Overall, respondents supported using national databases to investigate child outcomes, and said they would prefer for study findings to be communicated by HCP before treatment. A few participants raised concerns about data confidentiality.

### Strengths and limitations

This study’s strengths include its large UK-wide sample and comprehensive scope across short and long-term health and educational outcomes. This is in comparison to other qualitative surveys conducted on a similar population and setting (Boivin et al., 2020; Harrison et al., 2021; Sousa-Leite et al., 2023). The use of predominantly free-text questions yielded rich and diverse qualitative insights than structured questions allow (O’Cathain & Thomas, 2004), helping to address a gap in ART research that often prioritises clinical success over patient experience (Pennings & Ombelet, 2007).

Nonetheless, limitations should be considered when interpreting the results. Voluntary participation may have introduced self-selection bias (Bethlehem, 2010), with individuals who had more adverse experiences more likely to respond to the optional open-ended questions and thus potentially overrepresented (Redshaw et al., 2007). The predominantly White, highly educated sample may limit generalisability, though this distribution reflects patterns seen in the UK (Human Fertilisation and Embryology Authority, 2023) and other qualitative ART studies (Li et al., 2025; Mayette et al., 2024; Redshaw et al., 2007; Rothwell et al., 2020). Our UK-based findings may not be generalisable across other countries due to social and financial differences surrounding ART (Dancet et al., 2010). The cross-sectional design captures perceptions at a single time point and cannot determine how views may change throughout fertility treatment. Thematic analysis involves subjective interpretation, yet observer bias was minimised through researcher triangulation and peer debriefing (Nowell et al., 2017).

### Comparison with other studies

Respondents’ concerns about reproductive, neurological and developmental outcomes in their offspring are not unfounded. Berntsen et al. (2019) reviewed these domains and highlighted significant inconsistencies. For reproductive outcomes, some data suggest reduced sperm counts in ICSI-conceived male offspring, while no adverse effects were observed in female offspring (Berntsen et al., 2019). This potential sex-specific risk, however, is difficult for women to consider as the embryo sex is typically undisclosed during fertility treatment in the UK (Fox, 2009). Neurodevelopmental outcomes were similarly conflicted: associations with ART were often subgroup-dependent or disappeared after adjusting for multiple pregnancies (Berntsen et al., 2019). Furthermore, it is unclear whether these potential adverse outcomes stem from ART itself or the underlying infertility (‘chicken-or-egg dilemma’) (Berntsen et al., 2019; Graham et al., 2022). Such inconclusive evidence can be understandably difficult for women to disentangle, adding ambiguity and concerns to already complex fertility decisions, balancing personal hopes with fears for the child’s long-term health.

Our findings on the importance of communication of information align with previous research. Assaysh-Öberg et al.’s (2023) meta-ethnographic analysis of 19 qualitative studies highlighted women ‘felt uninformed about the long-term effects of treatment, on themselves and on their fetus and possible child’, and this lack of information can heighten their existing concerns. The literature has consistently emphasised the importance of providing high-quality information (Dancet et al., 2010) with transparency (Perrotta et al., 2025), sensitivity (Holter et al., 2021) and empathy by doctors (Malin et al., 2001), aligning with respondents’ requirements.

### Implications for clinical practice

Women’s concerns about long-term outcomes and suggestions for provision of information underpin the need for patient education and patient-centred communication. Apart from the top concerns identified in this survey, validated ART questionnaires such as the Concerns During Assisted Reproductive Technologies (CART) scale (Klonoff-Cohen & Natarajan, 2004) can inform clinicians about individual concerns and help tailor the content of information delivered. Information provision has been shown to reduce treatment-related concerns (Gameiro et al., 2013). Formalising this through online education programmes can improve psychological outcomes while offering a cost-effective solution for clinics (Cousineau et al., 2008). Overall, the literature suggests that patient education is both feasible and effective in addressing women’s concerns.

Yet, from providers’ perspectives, addressing women’s concerns amid inconclusive evidence can be challenging. Communication from HCP must balance transparency and sensitivity. National Institute for Health and Care Excellence (2013) clinical guidelines offer clear recommendations on discussing long-term ART safety, including areas where evidence is still awaited. Given the fear and anxiety expressed by respondents, providing emotional support from clinicians and psychologists (Dancet et al., 2011) may help navigate uncertainty and achieve patient-centred communication.

### Implications for future research

Women’s concerns about long-term outcomes, or the widespread lack of concern (reflecting limited information), can only strengthen the rationale for further national database studies, an approach broadly supported by respondents. Our research group will use these qualitative insights to inform our national data linkage study and may disseminate its findings to key stakeholders with consideration of respondents’ preferences, such as sharing results sensitively, via HCP and prior to treatment. As demonstrated in this survey and the wider literature, incorporating patient and public involvement (PPI) into ART studies can guide research priorities and build public trust. Trust is crucially important in fostering patient engagement with data use (Carson et al., 2019), and PPI has been shown to meaningfully impact the direction of laboratory (Fleming et al., 2021) and clinical research (Healey et al., 2024). Additionally, qualitative longitudinal studies with focus groups or interviews could provide deeper insights into how women’s views may evolve at different treatment stages. Future studies should also include ethnic minorities to address possible racial disparities in fertility research caused by cultural and systemic factors (Ekechi, 2021).

## Conclusion

Most women undergoing or considering ART reported no concerns or information about long-term health and educational outcomes for themselves or their children, yet some raised important concerns about maternal and offspring outcomes. Overall, participants supported using national databases to investigate these outcomes, highlighting the need for appropriate dissemination, patient-centred communication and PPI-informed research that reflects their requirements and concerns.

## Author contributions

**Faiza Afzal:** Conceptualization, Data curation, Formal analysis, Project administration, Writing – original draft, Writing – review & editing. **Ling Yin Fritz Wong:** Data curation, Formal analysis, Writing – original draft, Writing – review & editing. **Mitana Purkayastha:** Methodology, Validation, Writing – review & editing. **Yan Lu:** Validation, Writing – review & editing. **Philippa Rees:** Writing – review & editing. **Melissa A. Richard:** Writing – review & editing. **Carrie L. Williams:** Writing – review & editing. **Philip J. Lupo:** Writing – review & editing. **Barbara Luke:** Writing – review & editing. **Alastair G. Sutcliffe:** Conceptualization, Writing – original draft, Writing – review & editing.

## Acknowledgments

The authors thank all participants who responded to the survey and Fertility Network UK for facilitating its distribution.

All research at Great Ormond Street Hospital NHS Foundation Trust and UCL Great Ormond Street Institute of Child Health is made possible by the NIHR Great Ormond Street Hospital Biomedical Research Centre. The views expressed are those of the author(s) and not necessarily those of the NHS, the NIHR or the Department of Health.

## Disclosure of interest

The authors report there are no competing interests to declare.

## Additional information

### Funding

This work was supported by the Wellcome Trust (grant number: 2266971).

## Data availability statement

The data that support the findings of this study are available on request from the corresponding author, FA. The data are not publicly available as they contain information that could compromise the privacy of research participants.

## Notes

### Competing Interest Statement

The authors have declared no competing interest.

### Author Declarations

Ethical approval was obtained from University College Londons Research Ethics Committee (28371/001). A disclaimer was added to the participant information sheet with contact details of Fertility Network UK. This was to ensure that participants were provided adequate support if they were concerned or affected by any of the questions in the survey. All participants provided informed consent to participate in the research.

